# Can N95 respirators be reused after disinfection? And for how many times?

**DOI:** 10.1101/2020.04.01.20050443

**Authors:** Lei Liao, Wang Xiao, Mervin Zhao, Xuanze Yu, Haotian Wang, Qiqi Wang, Steven Chu, Yi Cui

**Affiliations:** 4C Air, Inc., Sunnyvale CA, USA; Department of Physics, Stanford University, Stanford CA, USA; Department of Molecular and Cellular Physiology, Stanford University, Stanford CA, USA; Department of Materials Science and Engineering, Stanford University, Stanford CA, USA; Stanford Institute for Materials and Energy Sciences, SLAC National Accelerator Laboratory, Menlo Park CA, USA

## Abstract

The Coronavirus Disease 2019 (COVID-19) pandemic has led to a major shortage of N95 respirators, which are essential to protecting healthcare professionals and the general public who may come into contact with the virus. Thus, it is essential to determine how we can reuse respirators and other personal protection in these urgent times. We investigated multiple commonly used and easily deployable, scalable disinfection schemes on media with particle filtration efficiency of 95%. Among these, heating (≤85 °C) under various humidities (≤100% RH) was the most promising, nondestructive method for the preservation of filtration properties in meltblown fabrics as well as N95-grade respirators. Heating can be applied up to 50 cycles (85 °C, 30% RH) without observation in the degradation of meltblown filtration performance. Ultraviolet (UV) irradiation was a secondary choice which was able to withstand 10 cycles of treatment and showed small degradation by 20 cycles. However, UV can also potentially impact the material strength and fit of respirators. Finally, treatments involving liquids and vapors require caution, as steam, alcohol, and household bleach may all lead to degradation of the filtration efficiency, leaving the user vulnerable to the viral aerosols.

---

COVID-19 is an ongoing pandemic with over a million confirmed cases and with new cases increasing by ∼10% per day (at the time of writing),^1^ that has caused major disruptions to nearly all facets of everyday life around the world. The disease is caused by the severe acute respiratory syndrome coronavirus 2 (SARS-CoV-2), which was first detected in Wuhan, China.^2,3^ The virus is of likely zoonotic origin, and like the SARS-CoV, enters human cells via the angiotensin-converting enzyme 2 (ACE2). ACE2 is a membrane protein that is an entry point for coronaviruses found in the lungs, heart, kidneys, and intestines that is responsible for regulating vasoconstriction and blood pressure. It was found that the SARS-CoV-2 utilizes the ACE2 more efficiently than SARS-CoV, which may explain why the human-to-human transmissibility of the virus is so high.^4^

Once infected, the patient will exhibit flu-like symptoms such as fever, chest tightness, dry cough, and in some cases development into severe pneumonia and acute respiratory distress syndrome (ARDS)^5–8^. As the incubation period is around 3-4 days but can be as long as 20 days, along with the presence of asymptomatic carriers, the virus has been extremely difficult to contain^9^. While the initial mortality rate was estimated to be around 3.5% in China, compounding the longer incubation period and testing delays has led to new global estimates of around 5.7%^10^.

While the exact mode SARS-CoV-2’s viral transmission is not known, a primary mode in viruses such as SARS and influenza is known to be through short-range aerosols and droplets.^11^ When a person infected with a virus breathes, speaks, sings, coughs, or sneezes micron sized aerosols containing the virus are released into the air. Data gathered from influenza patients suggest that these aerosols are typically fine (<5 μm) or coarse (>5 μm).^11–13^ For coarse particles, they can settle due to gravity within an hour. Fine particles, especially smaller than 1 μm, can essentially stay nearly indefinitely in the air. Droplets, or particles >10 μm, settle rapidly and are not typically deposited in the respiratory tract through means of aerosol inhalation. Particles >5 μm typically will only reach the upper respiratory tract, and fine particles <5 μm are critically able to reach the lower respiratory tract, similar to harmful particulate matter pollution (Figure 1). While coughing and sneezing do provide many aerosols, the size distribution and number of particles emitted during normal speech serve as a significant viral transmitter.^12^ Singing has been found to be comparable to continuous coughing in the transmission of airborne pathogens,^14^ which was demonstrated during a choir practice on March 10, 2020 in Washington State. Although the choir members did not touch each other or share music during the rehearsal, 45 out of the 60 members of the Skagit Valley Choir were diagnosed with the virus three weeks later, and two have deceased.

**Figure 1.**
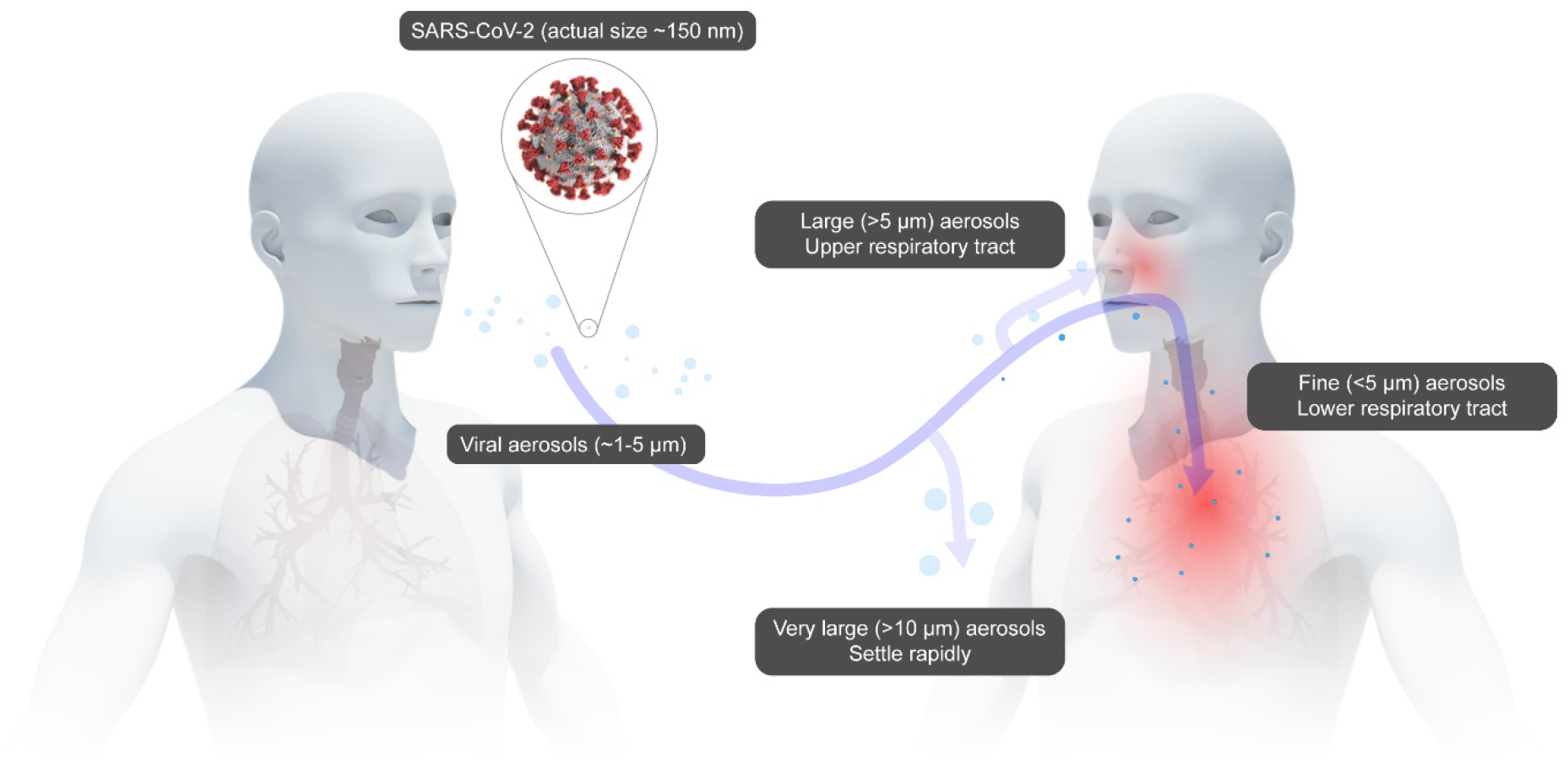
Transmission of SARS-CoV-2 through viral aerosols. Image of SARS-CoV-2 courtesy of the CDC.

For dangerous airborne particulates, including viral aerosols during the current COVID-19 pandemic, the United States Centers for Disease Control and Prevention (CDC) recommends the usage of N95 filtering facepiece respirators (FFR) as personal protective equipment for healthcare professionals.^15–17^ The N95 grade is determined by the CDC’s National Institute of Occupational Safety and Health (NIOSH) (document 42 CFR Part 84) which designates a minimum filtration efficiency of 95% for 0.3 μm (aerodynamic mass median diameter) of sodium chloride aerosols. In addition to N95, there are N99 and N100 which correspond to filtration efficiencies of 99% and 99.97%, respectively. For oil-based aerosols (DOP), NIOSH also has created grades R and P (with filtration efficiencies 95-99.97%). Elsewhere around the globe, the equivalent filtration grades to N95 are FFP2 (European Union), KN95 (China), DS/DL2 (Japan), and KF94 (South Korea). While the actual SARS-CoV-2 virus is around 150 nm,^18^ commonly found N95 respirators can offer protection against particles as small as 80 nm with 95% filtration efficiency (initial testing, not loaded).^19^ With the actual viral aerosols in the ∼1 μm range, the N95 FFRs’ filtration efficiency should be sufficient for personal protection.

The N95 FFR is comprised of multiple layers of, typically, polypropylene nonwoven fabrics (Fig. 2a).^20^ Among these layers, the most critical of which is that produced by the meltblown process. In typical FFRs, the meltblown layer is 100-1000 μm in thickness, comprised of polypropylene microfibers with a diameter in the range of ∼1-10 μm, as seen in the scanning electron microscope (SEM) images in Fig. 2b-c. Due to the production method, meltblown fibers produce a very lofty nonwoven where the fibers can stack and create a 3D network that has a porosity of 90%,^21^ leading to very high air permeability.

**Figure 2.**
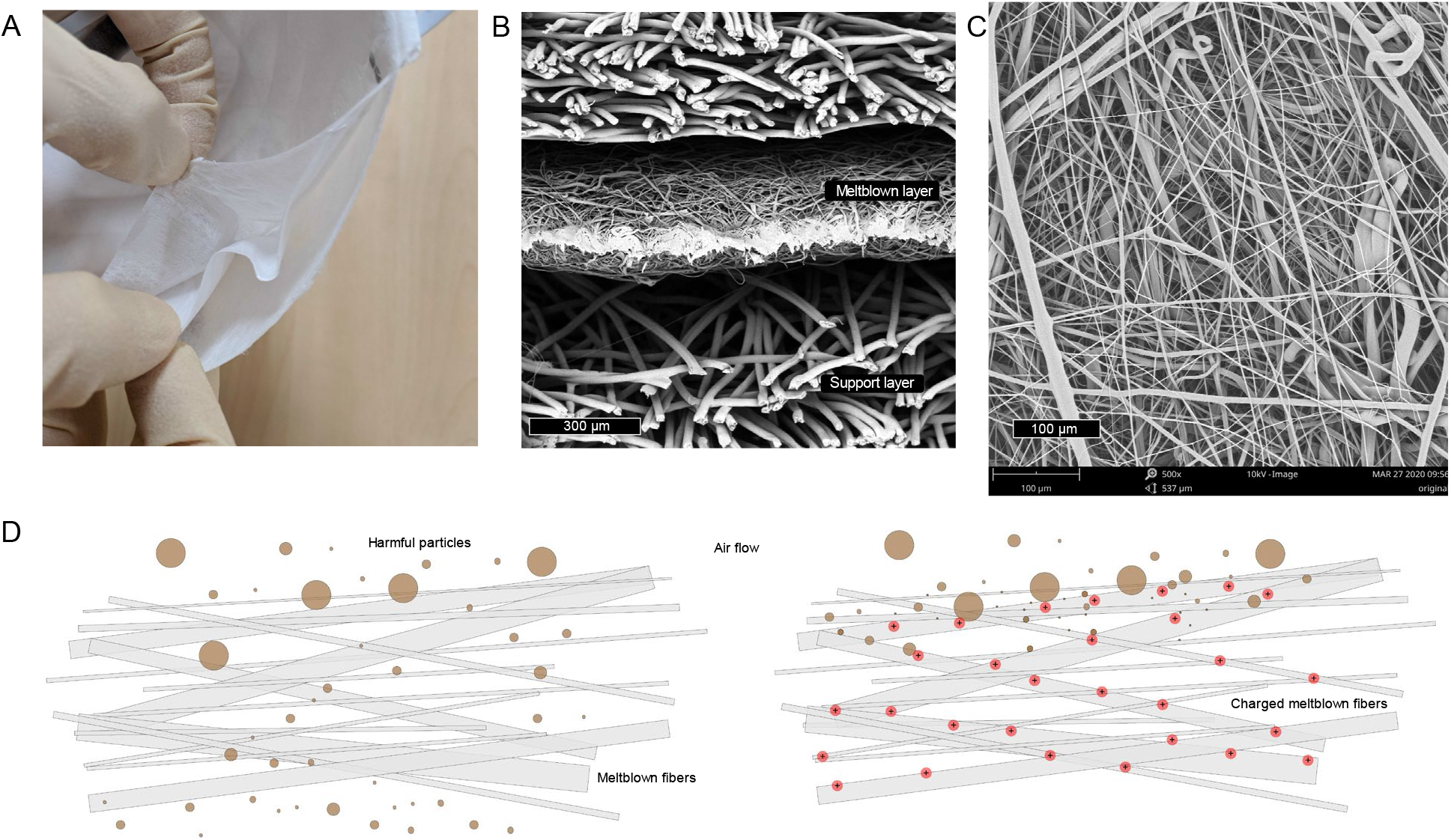
Meltblown fabrics in N95 FFRs A. Peeling apart a representative N95 FFR reveals multiple layers of nonwoven materials. B. SEM cross section image reveals the middle meltblown layer has thinner fibers with thickness around 300 μm. C. SEM image of meltblown fibers reveals a complicated randomly-oriented network of fibers, with diameters in the range of ∼1-10 μm. D. Schematic illustration of meltblown fibers without (left) and with (right) electret charging. In the left figure, smaller particles are able to pass through to the user, but particles are electrostatically captured in the case of an electret (right).

However, given that the fiber diameters are relatively small, and the filters’ void space is large, the filtration efficiencies of meltblown fabrics by themselves should not be adequate for fine particle filtration (Fig. 2d). To improve the filtration efficiency while keeping the same high air permeability, these fibers are charged through corona discharge and/or triboelectric means into quasi-permanent dipoles called electrets.^22,23^ Once they are charged, the filter can significantly increase its filtration efficiency without adding any mass or density to the structure (figure 2d). In addition, while other filter media may decrease in efficiency when loading the filter with more aerosol (NaCl, DOP), the meltblown electrets are able to keep a relatively consistent efficiency throughout the test.^24^

The COVID-19 pandemic has led to a significant shortage of N95 FFRs,^25^ especially amongst healthcare providers. Though the virus will eventually become inactive on the mask surface and it is unlikely to fully penetrate to the user’s intake side, a recent study shows that it required 72 hours for the concentration of SARS-CoV-1 and SARS-CoV-2 viruses on plastic surfaces (40% RH and 21-23 °C) to be reduced by 3 orders of magnitude (from 10^3.7^ to 10_0.6_ TCID_50_ per milliliter of medium).^26^ Assuming a similar longevity on FFR surfaces, it is important to develop procedures for the safe and frequent re-use of FFRs without reducing the filtration efficiency. The CDC has recommended many disinfection or sterilization methods, typically involving chemical, radiative, or temperature treatments.^27^ In brief, we can summarize the mechanism of disinfection or sterilization of bacteria and viruses as among the following: protein denaturation (alcohols, heat), DNA/RNA disruption (UV, peroxides, oxidizers), cellular disruption (phenolics, chlorides, aldehydes). While none of these methods have been extensively evaluated for SARS-CoV-2 inactivation specifically, we tested methods that can be easily deployed within a hospital setting with relatively high throughput for FFR reuse.

Among the CDC forms of disinfection, we chose five commonly used and potentially scalable, user-friendly methods: 1) heat under various humidities (heat denaturation inactivates SARS-CoV with temperatures >65 °C in solution, potentially the SARS-CoV-2 with temperatures >70 °C),^28–30^ 2) steam (100 °C heat based denature), 3) 75% alcohol (denaturing of the virus, based on the CDC), 4) household diluted chlorine-based solution (oxidative or chemical damage, based on the CDC), and 5) ultraviolet germicidal irradiation (UVGI was able to inactivate the SARS-CoV in solution with UV-C light at a fluence of ∼3.6 J/cm^2^).^28^ Using an oven, a UV-C sterilizer cabinet (found in barbershops or salons), steam, or liquid sprays can all realistically be deployed in the modern hospital setting and potentially in homes if needed. We did not consider some other common, but more inaccessible techniques (equipment such as electron beam irradiation or plasma generators can be expensive or dangerous), or some techniques known to cause damage to the FFR were not considered in our study.^31^

Due to the shortage of FFRs, data collected was tested on a meltblown fabric (20 g/m^2^) with initial efficiency ≥95% (full details are listed in the methods), unless otherwise specified. These samples are representative of how the filtration efficiency in a N95 FFR may change given exposure to these treatments in the worst-case scenario (i.e. no protective layer of the FFR). All meltblown samples were characterized using an industry standard Automated Filter Tester 8130A (TSI, Inc.) with a flow rate of 32 L/min and NaCl aerosol (0.26 μm mass median diameter). We subjected the meltblown samples to the aforementioned five disinfection methods and summarized the data in Table 1.

**Table 1.** One-time disinfection treatment on a meltblown fabric

| Treatment | Mode of application | Treatment time (min) | Filtration efficiency (%) | Pressure drop (Pa) |
| --- | --- | --- | --- | --- |
| Initial samples | | | 96.52 ± 1.37 | 8.7 ± 1.0 |
| Dry heat (75 °C) | Static-air oven | 30 | 96.21 | 7.00 |
| Steam | Beaker of boiling water | 10 | 94.74 | 8.00 |
| Ethanol (75%) | Immersion and air dry | Until dry | 56.33 | 7.67 |
| Chlorine-based (2%) | Light spray and air dry | 5 | 73.11 | 9.00 |
| UVGI (254 nm, 8 W) | Sterilization cabinet | 30 | 95.50 | 7.00 |

From the first disinfection, we can clearly note that the solution-based methods (ethanol and chlorine-based solution) drastically degraded the filtration efficiency to unacceptable levels, while the pressure drop remained comparable. As the pressure drop remained constant, this indicated that the loftiness and structure of the meltblown was unchanged, and the resultant efficiency degradation is the result of depolarization from the quasi-permanent polarization state of the electret (Figure S1). It is hypothesized in the literature that small molecules such as solvents can permeate into the fabric and liberate the charge traps or frozen charges of the electret,^32^ which would decrease the filtration efficiency. It is also possible that the chlorine-based solution may also degrade the efficiency less than the alcohol-based solution due to the higher water content. As polypropylene is hydrophobic, the chlorine-based solution may have a more difficult time in the penetration into the fabric and the static charge of fibers deeper within the meltblown may be less affected.

These initial results are mostly in agreement with a NIOSH published report regarding the decontamination of whole FFRs,^33^ though it placed more focus on gas-based methods, which could be suitable to well-controlled industrial scale disinfection. However, this may not be practical for on-site disinfection within the current hospital and clinic infrastructure. They found that bleach (immersion in a diluted solution) resulted in a less of an efficiency drop than our results, but noted there were strong odors from off-gassing, which is another reason to exclude this as a method to consider for the end-user.

We chose to focus on the three remaining treatment methods to perform multiple treatment cycles. The meltblown fabrics after ten cycles of each treatment are summarized in Figure 3 (data provided in Table S1). After three treatments of these three methods, the meltblown fabric still has characteristics similar to the initial sample. However, after five steam treatments, the efficiency has a sharp drop which continues at cycle 10 (Table S2). Similar to the alcohol and chlorine solution treatments, the pressure drop can be maintained at ∼8-9 Pa, but the efficiency has degraded to around ∼80%, which would be concerning in an environment with high viral aerosol concentrations. As with the solution treatments, the pressure drops remained similar, which suggests it is also due to the decay of static charge. The dwelling time and frequency may be critical for how well the static charge can be preserved. If steam treatments saturate the fibers many times and can condense water droplets on the fibers, it is possible that the static charge decays after multiple treatments. As this decay is due to the direct water molecule contact with the fibers, it may be possible to alleviate the static decay if the fibers do not come into contact with the vapor directly (sealed container, apparatus, bag, etc.) and steam only serves as the heating element.

**Figure 3.**
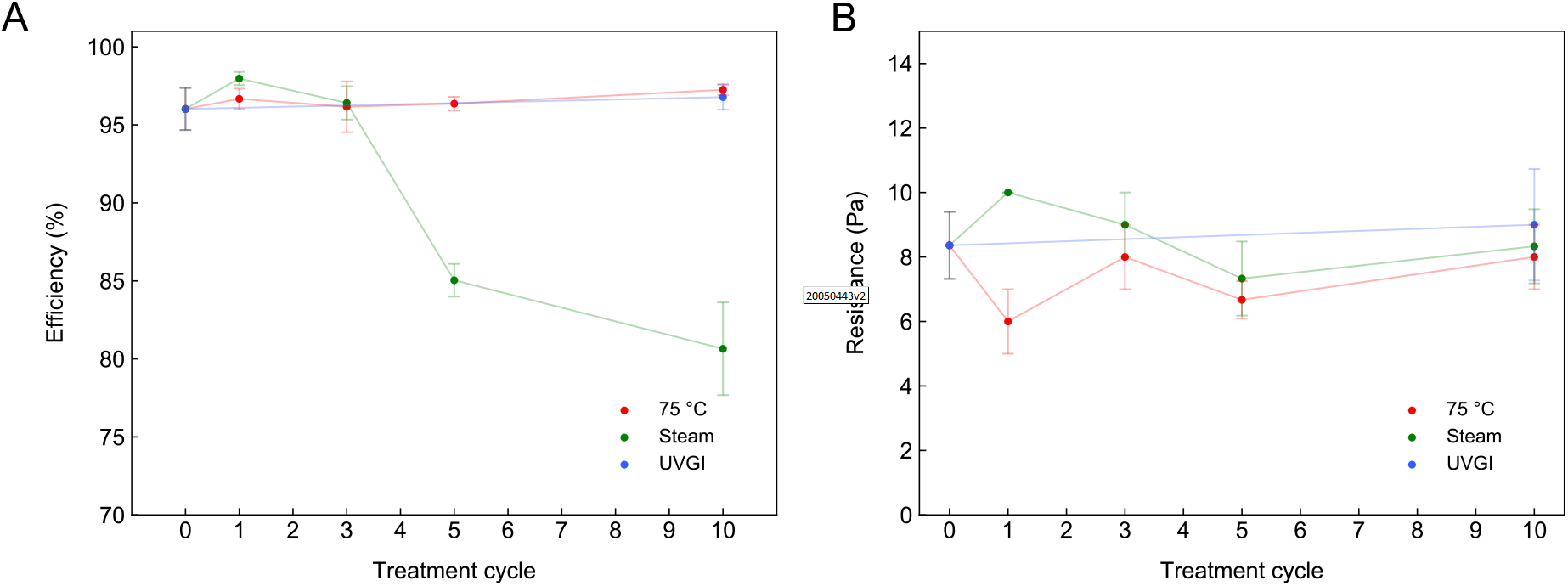
Ten treatment cycle evolution of filtration characteristics. A. Efficiency evolution where it is clear that steam treatment results in a degradation of efficiency. B. Pressure drop evolution where it is not apparent that any structure or morphology change has occurred in the meltblown fabrics.

Given that steam also resulted in eventual efficiency degradation, we further determined the limits of temperature and humidity. We performed multiple humidity experiments (30%, 70%, and 100% RH) at 85 °C (20 minutes/cycle), observing that no appreciable degradation of efficiency at any humidity level (Fig. 4a-b). At 85 °C, 30% RH, we observed no efficiency degradation over fifty cycles on a meltblown fabric (Fig. 4c-d). Using less harsh conditions (75 °C, dry heat), the results are expectedly in agreement (Figure S2). This is further confirmed when testing multiple N95-level FFRs from various countries (listed in methods) at 85 °C, 30% and 100% RH for 20 cycles (Fig. 4e-f). Testing conditions for the FFRs were under a flow rate of 85 L/min. From all the FFRs, we observed little change in the filtration properties, as all FFRs with filtration efficiency >95% were able to retain filtration efficiencies >95% after 20 cycles of heat treatment, even in a humid environment.

**Figure 4.**
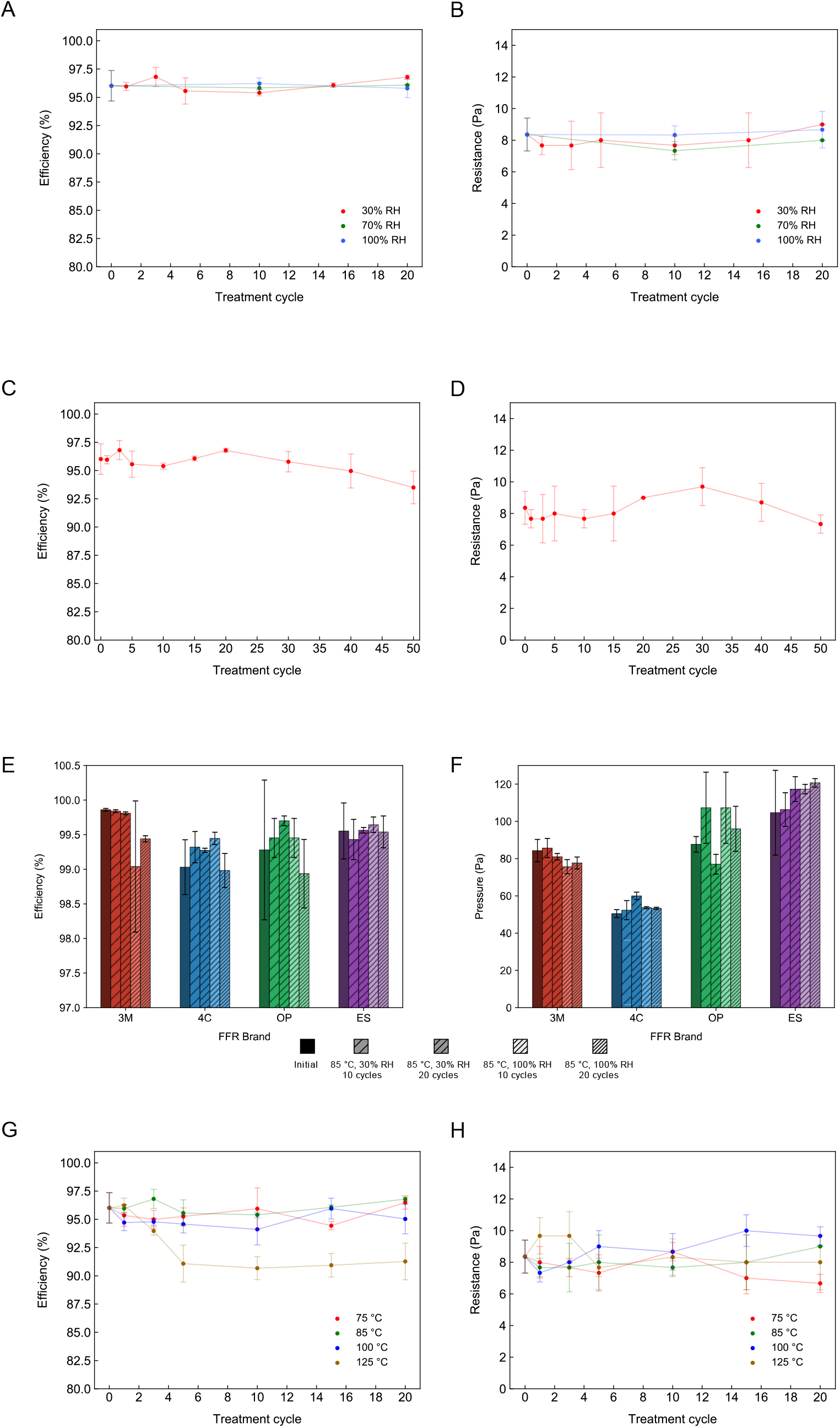
Temperature and humidity evolution of meltblown and FFR filtration characteristics. A-B. Evolution of filtration characteristics at 85 °C under different humidities, efficiency (A) and pressure drop (B). C-D. Evolution of filtration characteristics on a meltblown fabric under 85 °C, 30% RH, efficiency (C) and pressure drop (D). E-F. Evolution of the filtration characteristics on an N95-level FFRs with 85 °C, under 30% and 100% RH (measured at a flow rate of 85 L/min), efficiency (E) and pressure drop (F). The left-to-right of all FFR brands is as follows: 1) initial (leftmost, solid pattern, tested in ambient conditions), 2) 85 °C, 30% RH 10 cycles, 3) 85 °C, 30% RH 20 cycles, 4) 85 °C, 100% RH 10 cycles, 5) 85 °C, 100% RH 20 cycles. G-H. Temperature dependence of filtration characteristics over 20 cycles with RH<30%, efficiency (G) and pressure drop (H).

To determine the upper limit of applicable temperature, we tested low humidity conditions (≤30% RH) up to 125 °C (10 minutes/cycle), plotted in Fig. 4g-h (data provided in Table S3). There is little to no change in the filtration efficiency and pressure drop up to 100 °C in low moisture conditions. However, at 125 °C, there is a sharp drop in the filtration efficiency while maintaining a constant pressure drop at around cycle 5. Similarly, the lack of pressure drop change indicates that the degradation is also due to the static decay. Considering the higher temperature, polypropylene’s melting point (130-170 °C), as well as the thin and fibrous nature of the media, it is possible that the higher temperature is enough to relax the microscopic charge state within the polymer resulting in some of the quasi-stable polarization to become depolarized to their neutral state. From SEM images, we did not identify any morphological changes and did not observe any apparent physical deformations (Figure S3), which may support this conclusion. This effect is not as strong as direct solvent contact, which reduced the filtration efficiency to <80%, while 125 °C reduced the efficiency to ∼90%. The mechanism of depolarization may differ, as the solvent can liberate charge traps, but the temperature here provides energy to return some of the fibers’ polarized state to a relaxed state.

Heat was a very promising scalable method that may be suitable for FFR reuse. We can conclude that the highest subjectable temperature to the FFR for repeated use with ≥95% efficiency is <100 °C. At temperatures ≤85 °C, humidity does not seem to play a crucial role in the filtration properties, as FFRs tested at a near 100% RH at 85 °C were unaffected. However, as steam results in a decrease in efficiency, the humidity should be kept low if approaching 100 °C. The temperature range here may pose some limitations in the available equipment, but we believe the current hospital infrastructure should be able to perform these treatments. This includes using dryers, ovens, circulators, or even hot air guns, all of which are relatively scalable and user-friendly. Repeated treatments at temperatures below 65 °C (30 minutes) is not advised, as it was the reported temperature required to inactivate the SARS-CoV in solution and limit it to undetectable traces.^29^ As the prior inactivation tests occurred in solution, there requires further study to be done on the heat inactivation of aerosolized viruses.

Finally, we tested the effect of UVGI on meltblown samples up to twenty cycles (Figure 5). Fig. 5. The UVGI sterilization cabinet here provides UV-C light centered at 254 nm with intensity of 8 W. The UV-C light areal intensity distribution is not uniform inside the cabinet and its exact value needs to be measured in the future for dose determination, as the necessary radiation to inactivate SARS-CoV was previously found to be above ∼3.6 J/cm^2^.^28^ At ten cycles, the data is in agreement with the NIOSH report,^33^ but eventually decays to 93% at twenty cycles and makes it unsuitable for N95-grade FFRs by itself.

**Figure 5.**
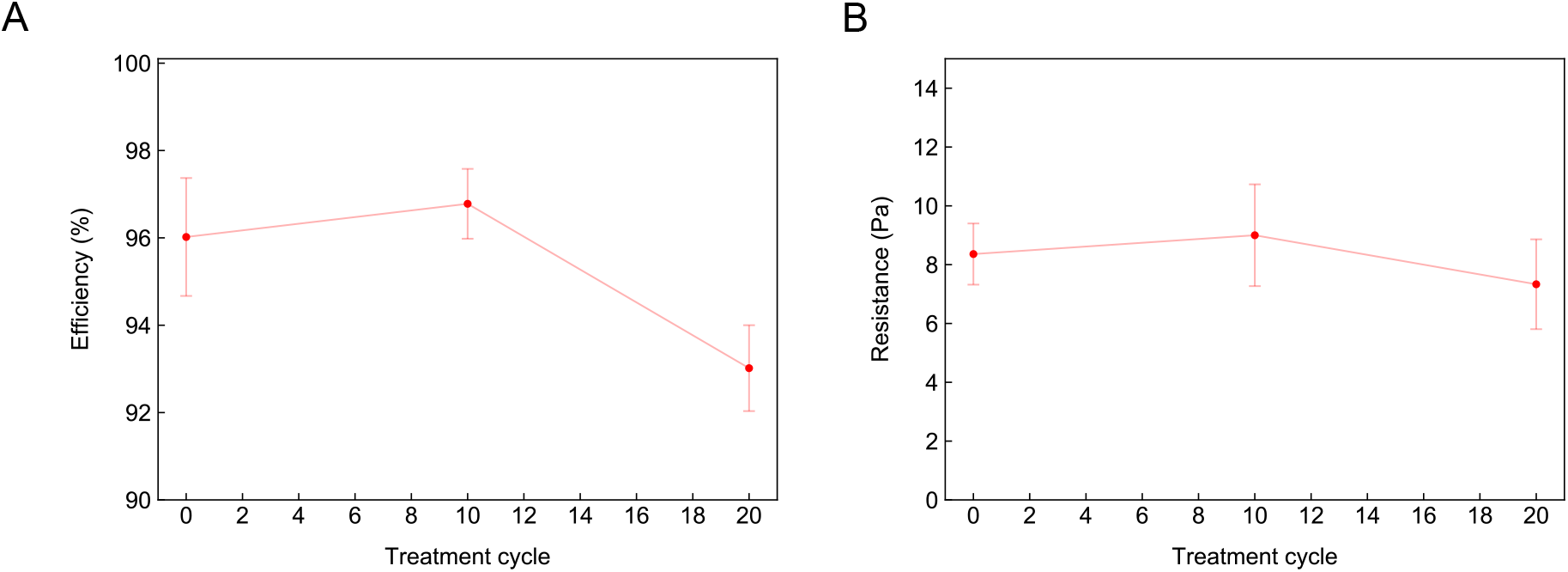
Effect of UVGI on meltblown filtration characteristics. A. Efficiency of meltblown fabric that slightly changes after 10 cycles of UVGI. B. Pressure drop after UVGI treatments remains similar

While UV radiation may possess enough energy to break the chemical bonds and degrade polypropylene, the dosage of the sterilization chamber is relatively low, and the material degrades very slowly. This is supported by previous experiments that showed UV-C doses up to 950 J/cm^2^ did not appreciably change the filtration efficiency.^34^ A possible concern regarding UVGI disinfection for FFRs is of the UV penetration depth. As UV-C has a wavelength around 250 nm, and polypropylene is a UV absorber, it is difficult to conclude if smaller viral particles deep within the filter can be deactivated through UVGI. If the particles are of a larger size and remain localized on the surface, UVGI may be a candidate for FFR reuse. Furthermore, this means that UVGI requires FFRs to not be stacked, as the incident radiation is will only be absorbed by the top-most surface. Another disadvantage is that UVGI was reported to significantly impact the mechanical strength of some FFRs with doses of around 1000 J/cm^2^.^34^

Therefore, UVGI may be a useful disinfection technique, but the exact exposure or intensity of the UV-C light fluence on the mask surface would need to be verified. The variation in UVGI intensity has been the cause of discrepancies in the literature, as 3M’s own internal reports recently showed that their UVGI treatments damaged particular FFRs,^35^ whereas other reports show that UVGI cycling on multiple N95 FFRs had minimal or no impact.^31^

While most of our tests were performed on meltblown fabrics with initial efficiency >95% due to the current shortage of FFRs, there is a definite concern over whether other FFR components (straps, valves, nosepiece, foam, etc.) can change in these treatment environments. These can impact the fit and sealing of the FFR, which is equally important as the FFR efficiency itself. From our experiments, we also used typical N95-grade FFRs to test the strap elasticity and structural integrity after heat treatments. We noted no apparent or qualitative change in the strap elasticity or fit compared with the untreated model for the heat treatments, but future quantitative studies need confirm this.

As actually donning and using the respirator impacts the structural stability of such components, particular care needs to be taken in interpretation of these results. Previous reports also indicated that FFRs can safely be donned up to five times, but beyond five times may result in a less than adequate fit.^36^ During this crisis, users have to make sure that the fit of the FFRs after treatment are adequate and are not left vulnerable due to leakage. This may require mask producers or users to consider straps which are more robust for reuse or can withstand treatment conditions.

In conclusion, COVID-19 is an extremely contagious disease that requires healthcare professionals to take caution with necessary protective equipment. The current shortage of N95 FFRs during this time of rapidly spreading infection may be mitigated by methods that will allow the safe reuse. We have tested methods which may be suitable for the re-use of particulate respirators, and hope our results will be useful in helping hospitals and other health care facilities in formulating safe standard operating procedures (SOPs) so that virus inactivation is assured while not compromising mask protection. While we reiterate that these methods have not been tested on SARS-CoV-2 directly for inactivation, these methods use precedents set by either SARS-CoV or general disinfection strategies. We found that of commonly deployable methods, heating (dry or in the presence humidity) <100 °C can preserve the filtration characteristics of a pristine N95 respirator. The UVGI (254 nm, 8 W) sterilizer cabinet used in these tests does not have enough dose to damage the filtration properties within a reasonable number of treatment cycles and may be considered for disinfection, however the exact dose output of the cabinet would need to be determined such that it is suitable for inactivation of SARS-CoV-2 with minimal FFR damage. Using steam to disinfect requires caution, as the treatments may seem to be suitable, but prolonged treatment may leave the user with unsuitable protection. Finally, we advise against using certain liquid contact, such as alcohol solutions, chlorine-based solutions, or soaps to clean the respirator, as this will lead to a degradation in the static charge that is necessary for the FFR to meet the N95 standard.

## Sample preparation

A 20 g/m^2^ meltblown fabric with initial filtration efficiency of ≥95% was used for all samples. Each sample was cut to approximately 15 cm × 15 cm. All sample testing was performed on an Automated Filter Tester 8130A (TSI, Inc.) using a flow rate of 32 L/min and NaCl as the aerosol (0.26 μm mass median diameter). Each average measurement contains at least 3 individual sample measurements.

We chose disposable N95-grade FFRs for testing: 3M 8210 (NIOSH N95), 4C Air, Inc. (GB2626 KN95), ESound (GB2626 KN95) and Onnuriplan (KFDA KF94). These FFRs are referred to as “3M”, “4C”, “ES”, and “OP”, respectively in figures. Full FFR testing used a flow rate of 85 L/min.

SEM images were recorded using a Phenom Pro SEM, at 10 kV.

## Heat treatment

Samples were loaded into a pre-heated 5-sided heating chamber (Across International, LLC or SH-642, ESPEC) for the temperatures and times given in the main text. Dry heat was applied using the Across International vacuum heating oven under ambient conditions. In the case of the SH-642, the humidity was set the lowest value (30% RH up to 85 °C, above 85 °C the humidity is <30% but cannot be controlled). High humidity (100% RH) was simulated via sealing meltblown fabrics, or FFRs, inside a polyethylene bag with 0.3 mL of water and placing them inside the SH-642 chamber. The resting time between cycles was 10 minutes for the 75 °C and 85 °C treatments and 5 minutes for the 100 °C and 125 °C treatments. After resting, the samples were returned to the chamber to begin the next cycle. We initially chose 75 °C due to the presence of blanket warming ovens in hospital environments that can reach ∼80 °C. Further experiments used 85 °C, in the event that 75 °C is not enough to inactivate SARS-CoV-2. Microwaving was not considered as many FFRs contain metals which may spark and melt the fabric.

## Steam treatment

Three samples were stacked on top of a beaker with boiling water inside (at around 15 cm above the water). The samples were left on top of the beaker and steamed for ten minutes, afterwards they were left to air dry completely (to touch). Samples were either tested or placed back on top of the beaker to continue the next treatment cycle.

## Alcohol treatment

Samples were immersed into a solution of 75% ethanol and left to air dry (hanging) and subsequently tested.

## Chlorine-solution treatment

Samples were sprayed with approximately 0.3-0.5 mL of household chlorine-based disinfectant (∼2% NaClO). Samples were left to air dry and off-gas completely, hanging. Samples were tested.

## UVGI

Samples were placed into a UV sterilizer cabinet (CHS-208A), with a 254 nm, 8 W lamp, and 475 cm^2^ internal area. Samples were irradiated for 30 minutes and let to stand under ambient conditions for 10 minutes per cycle. Samples were either returned to the chamber for the next cycle or tested.

## Data Availability

All relevant data is included in the manuscript text and/or included supplementary materials

## Acknowledgements

We would like to thank Dr. Larry Chu and Dr. Amy Price at Stanford Health Care for the helpful discussion.

